# Immunothrombotic Features of Coronary Thrombi in Myocardial Infarction after SARS-CoV-2 Vaccination

**DOI:** 10.64898/2026.08.04.26359712

**Authors:** Ana Blasco, Beatriz Pelacho, María-José Coronado, Ana Royuela, Paloma Martín, Andrea Matutano, Ariana Castellano, Juan M. Escudier, Carolina González-Andrés, Javier Ortega, Carmen Bellas

**Author notes:** **Address for correspondence** Ana Blasco, MD, Cardiology Department, Hospital Universitario Puerta de Hierro-Majadahonda, C/Manuel de Falla 1, 28222 Madrid, Spain;).

## Abstract

**Background:** Neutrophil extracellular traps (NETs) contribute to immunothrombosis and arterial thrombosis. Mechanisms underlying myocardial infarction after SARS-CoV-2 vaccination remain poorly understood.

**Objectives:** To investigate histopathologic and immunothrombotic features of coronary thrombi in patients with ST-elevation myocardial infarction (STEMI) after SARS-CoV-2 vaccination.

**Methods:** We performed a retrospective matched cohort study including patients with STEMI undergoing primary percutaneous coronary intervention between January 2021 and March 2023. Coronary thrombi obtained by aspiration were analyzed by histopathology, immunohistochemistry, and confocal microscopy for NET detection. Vaccinated and unvaccinated patients were matched by age and sex. Associations between vaccination status and thrombus characteristics were assessed after adjustment for SARS-CoV-2 serologic status.

**Results:** Among 44 matched patients (23 vaccinated and 21 unvaccinated), NETs were identified in 14 vaccinated patients (61%) and 5 unvaccinated patients (24%; P = .01). Vaccination was associated with increased odds of NET-positive thrombi after adjustment for SARS-CoV-2 serology (odds ratio, 5.1; 95% CI, 1.36–19.45; P = .02). No associations were observed between vaccination and polymorphonuclear cell density, fibrin deposits, plaque fragments, or anti–platelet factor 4 staining. Among patients vaccinated within 100 days before STEMI, NET-positive thrombi were associated with shorter intervals between vaccination and myocardial infarction (median [IQR], 25 [11–64] vs 57 [40–84] days; P = .02).

**Conclusions:** SARS-CoV-2 vaccination was associated with increased NET presence in coronary thrombi from patients with STEMI, suggesting a potential NET-mediated immunothrombotic mechanism independent of classical vaccine-induced immune thrombotic thrombocytopenia.

## Introduction

Neutrophil extracellular traps (NETs) are key mediators of immunothrombosis and have been implicated in mechanical vessel obstruction during SARS-CoV-2 infection^1^, including coronary thrombosis leading to ST-elevation myocardial infarction (STEMI)^2^. Observational studies and case reports have described myocardial infarction temporally associated with COVID-19 vaccination^3–5^. However, large population-based analyses have not shown a consistent increase in myocardial infarction risk after vaccination, and available evidence does not support a definitive causal association^6,7^. In contrast, the risk of serious hematologic and vascular events appears substantially higher and more prolonged after SARS-CoV-2 infection than after vaccination^8^.

Vaccine-induced immune thrombotic thrombocytopenia (VITT) is a rare but well-characterized complication, defined by thrombocytopenia, markedly elevated D-dimer levels, anti–platelet factor 4 (PF4) antibodies, and thrombosis —often at atypical sites— with an estimated incidence of 3-10 cases per million doses^9^. Anti-PF4 antibodies generated after adenoviral vector vaccination can activate platelets and neutrophils, promoting NET formation and thrombosis^10^. Beyond VITT, immune activation and potential autoantibody responses following vaccination remain incompletely understood, complicating causal inference.

We investigated histopathologic mechanisms of coronary thrombosis in patients presenting with acute myocardial infarction after SARS-CoV-2 vaccination.

## Methods

### Ethical Approval

The study complied with the Declaration of Helsinki and was approved by the institutional ethics committee. Written informed consent was obtained.

### Study Design and Population

This retrospective cohort study included consecutive patients with STEMI who underwent primary percutaneous coronary intervention (PPCI) at a tertiary academic center between January 1, 2021, and March 9, 2023. All patients provided informed consent. Among 465 treated patients, thrombus aspiration was performed in those with large culprit arteries and high thrombus burden according to guideline recommendations. Intracoronary thrombi were obtained from 116 patients. Vaccination data were available for all 116 patients and SARS-CoV-2 serological data for 112.

Vaccinated patients were matched 1:1 to unvaccinated patients by age (±3 years) and sex (except in two cases matched 2:1), resulting in 21 matched sets (n = 44; 23 vaccinated and 21 unvaccinated). In a secondary analysis, all patients vaccinated within 100 days before myocardial infarction (n = 32) were evaluated for an association between time since vaccination and the presence of NETs (Figure 1).

**Figure 1.**
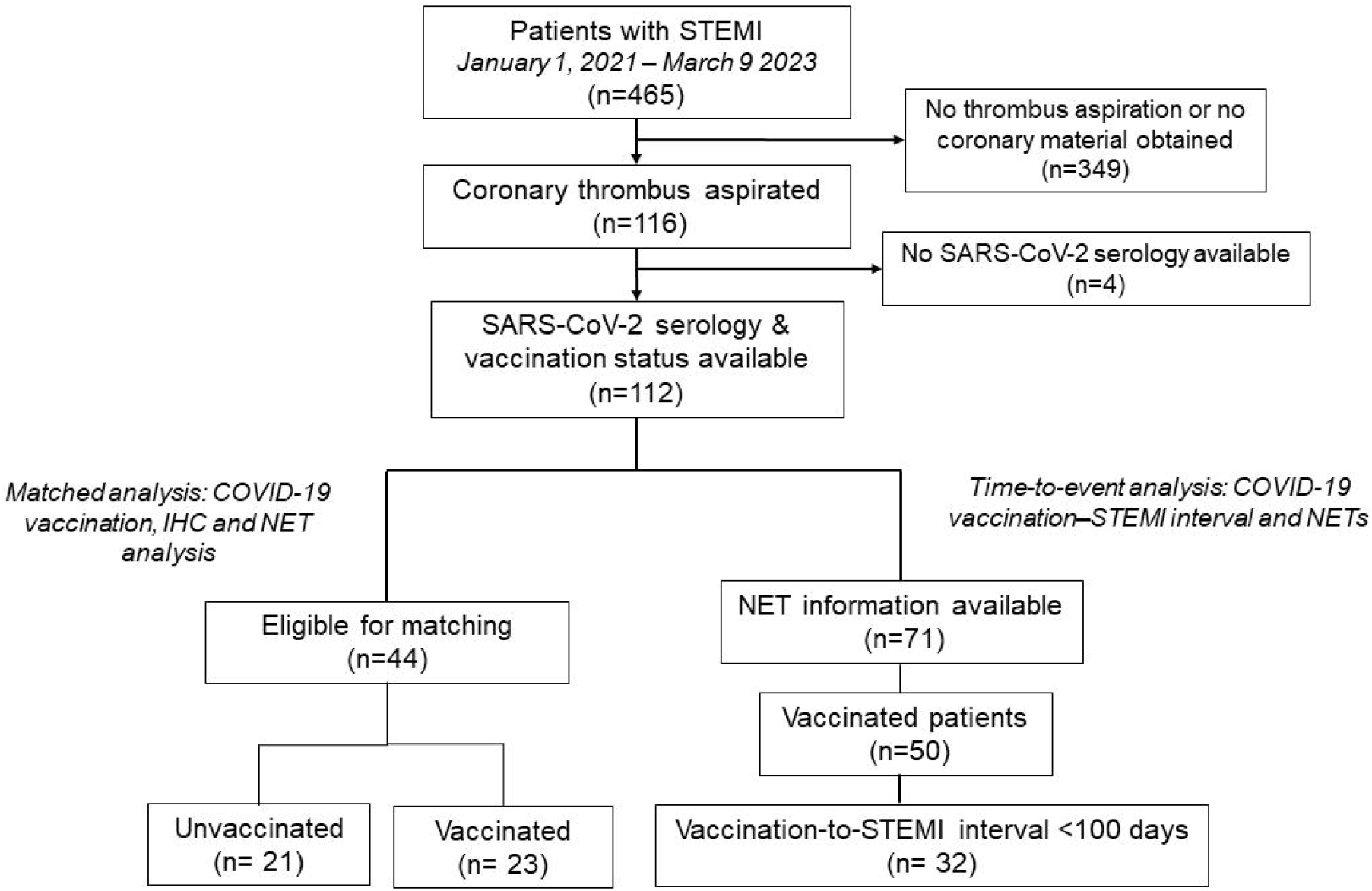
Flowchart of patients. Data are presented for all patients assessed for eligibility and included in the final analysis. Exclusion criteria and reasons for exclusion are detailed in the flow diagram. Abbreviations: IHC, immunohistochemical; NET, neutrophil extracellular traps; STEMI, ST-elevation myocardial infarction.

### Histopathologic and NET Analysis

Thrombi were formalin-fixed, paraffin-embedded, and stained with hematoxylin-eosin. Immunohistochemistry (IHC) was performed using antibodies against CD68, myeloperoxidase [Dako, Glostrup, Denmark] and PF4 [Bio-Techne R&D Systems]. NETs were identified by confocal microscopy based on colocalization of myeloperoxidase–DNA complexes and citrullinated histone 3^11^ (Figure 2). Quantification used image analysis software with Pearson correlation coefficients.

**Figure 2.**
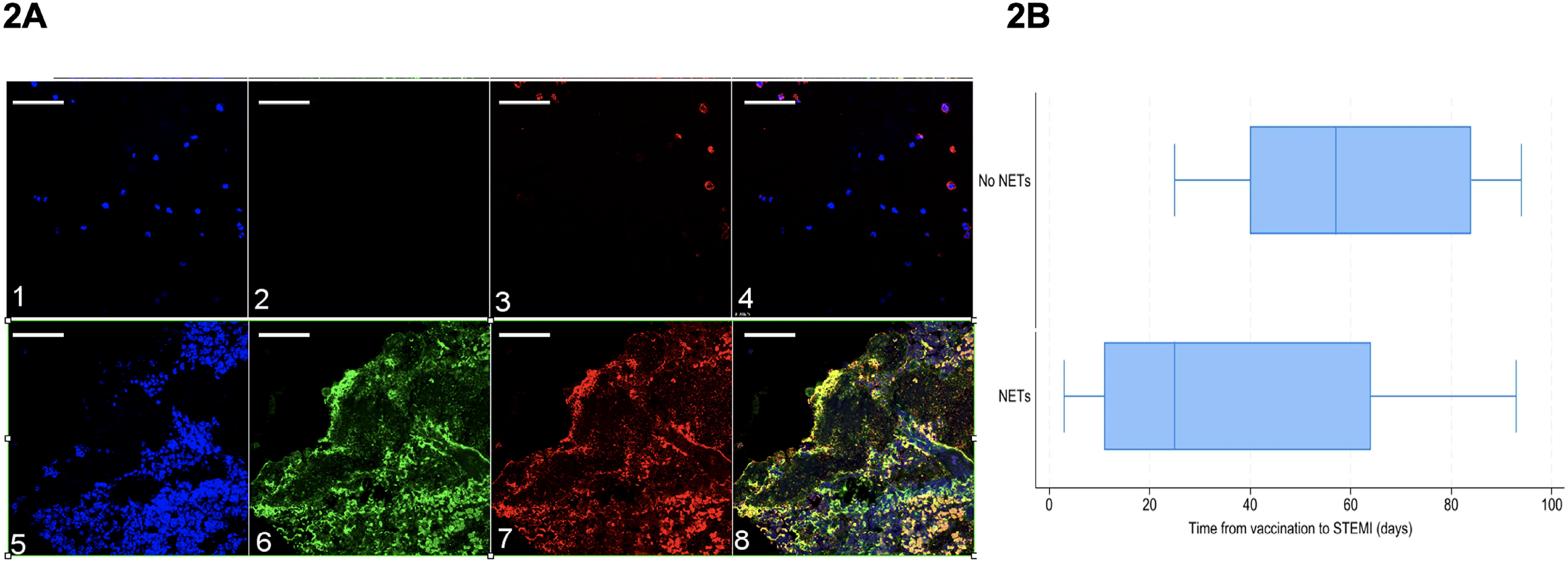
2A, Immunofluorescence of Neutrophil Extracellular Traps in Thrombi Aspirated During Primary Coronary Intervention. 2B, Time-to-event Analysis from COVID-19 vaccination to STEMI and NET formation. **2A.** (1–4) Representative images of coronary aspirate from a patient with ST-elevation myocardial infarction (STEMI) without neutrophil extracellular traps (NETs). (5–8) Representative images of coronary aspirate from a patient with STEMI with NETs. (1, 5) DNA stained with TO-PRO to identify nuclei; (2, 6) Neutrophils stained with a myeloperoxidase-specific antibody; (3, 7) Samples stained with a citrullinated histone H3–specific antibody; (4, 8) Merger of the three labels. Scale bars: 50 μm. **2B**. Box-and-whisker plots show the distribution of time from vaccination to STEMI among patients who experienced myocardial infarction within 100 days after vaccination, stratified by the presence of NETs in coronary thrombi. The centerline indicates the median; the box limits, the interquartile range (IQR); and the whiskers, the minimum and maximum values. Median (IQR) time was 25 (11–64) days in patients with NETs vs 57 (40–84) days in patients without NETs (P = .02). Abbreviations: NET, neutrophil extracellular traps; STEMI, ST elevation myocardial infarction

### SARS CoV-2 serologic testing

On admission, total antibodies (IgM, IgA, IgG) against nucleocapsid antigens were measured by electrochemiluminescence immunoassay [Elecsys Anti-SARS-CoV-2, ROCHE]. In positive cases, IgM and IgG antibodies against spike antigens were assessed by chemiluminescence immunoassay [LIAISON SARS-CoV-2, DIASORIN]; with IgG quantified within a detection range of 13.0–2,080 AU/mL.

### Statistical Analysis

Continuous variables are presented as mean (SD) or median (IQR), and categorical variables as number (percentage). Comparisons used Mann-Whitney U or χ^2^ tests. In the matched cohort, associations between vaccination and thrombus features were evaluated using conditional logistic regression adjusted for SARS-CoV-2 serologic status. Odds ratios (ORs) and 95% CIs were estimated. Associations between NET presence and time since vaccination were assessed using the Mann-Whitney U test.

## Results

Among 112 eligible patients, 44 were included in the matched analysis (38 men [86%]; mean [SD] age, 59 [12.5] years). Clinical and angiographic characteristics were similar between groups, except for peak troponin I level, which was higher in vaccinated patients (median 4,275 vs 274 ng/L; P = .03) (Table).

NETs were identified in 14 of 23 vaccinated patients (61%) and 5 of 21 unvaccinated patients (24%) (P = .01). After adjustment for SARS-CoV-2 serologic status, vaccination was associated with higher odds of NET presence in coronary thrombi (OR, 5.1; 95% CI, 1.36–19.45; P = .02). Vaccination was not associated with the density of polymorphonuclear cells, fibrin deposits, plaque fragments, or anti-PF4 staining.

NET analysis was available for 72 of 112 patients; 50 had received vaccination. Among vaccinated patients, 42 (84%) received mRNA vaccines and 8 (16%) received adenoviral vector vaccines. NETs were identified in thrombi from 26 of 42 patients (62%) who received mRNA vaccines compared with 4 of 8 patients (50%) who received adenoviral vector vaccines (P = .70). The interval between the most recent vaccine dose and STEMI was shorter in patients with NET-positive thrombi than in those without detectable NETs (median [IQR], 57 [17–124] vs 92 [53–406] days; P = .02).

To avoid potential selection bias due to the wide time interval between vaccination and STEMI, the analysis was repeated in all consecutive patients who experienced myocardial infarction within 100 days after vaccination (n = 32), and the association persisted (median [IQR], 25 [11-64] vs 57 [40-84] days; P = .02) (Figure 2).

The proportion of patients with detectable NETs varied according to the number of vaccine doses received: NETs were observed in 57.1% (4/7), 57.7% (15/26), 71.4% (10/14), 0% (0/2), and 100% (1/1) of patients receiving 1, 2, 3, 4, and 5 doses, respectively.

## Discussion

In this cohort of patients with STEMI, SARS-CoV-2 vaccination was associated with increased NET formation in coronary thrombi and shorter vaccination-to-event intervals among NET-positive cases. No associations were observed with other histologic thrombus features or anti-PF4 staining. These findings suggest a selective NET-mediated immunothrombotic mechanism associated with SARS-CoV-2 vaccination independent of the VITT.

NETs contribute to arterial thrombosis by providing a scaffold for platelets and coagulation factors and amplifying inflammatory and procoagulant pathways^12^. During SARS-CoV-2 infection, excessive NETosis has been implicated in endothelial injury and coronary thrombosis^1,2^. Our observations raise the possibility that similar immunothrombotic mechanisms may be present in a subset of post-vaccination coronary events.

VITT represents a distinct immune-mediated syndrome characterized by anti-PF4 antibodies, platelet activation, neutrophil stimulation, and NET release^9,10^. However, patients in this study did not exhibit thrombocytopenia or histopathologic evidence consistent with VITT, and anti-PF4 staining was not associated with vaccination. Most vaccinated patients received mRNA vaccines, whereas classical VITT has been linked predominantly to adenoviral vector platforms. These findings argue against a canonical PF4-dependent mechanism.

Alternative pathways may be relevant. Complement activation has been proposed as an intrinsic feature of mRNA–lipid nanoparticle platforms and may contribute to inflammatory adverse events. Complement-mediated platelet activation and neutrophil priming could theoretically lower the threshold for NET formation in susceptible individuals. Additionally, transient increases in NETosis biomarkers and autoantibodies capable of promoting NET formation have been described following vaccination^13^. Nonetheless, experimental evidence suggests that the spike protein alone does not directly induce robust neutrophil activation or NETosis^14^, suggesting the implication of host-specific immune responses.

The observed temporal gradient —characterized by shorter vaccination-to-STEMI intervals in NET-positive cases— supports biological plausibility. However, thrombotic risk is substantially higher following SARS-CoV-2 infection than after vaccination^8^. Moreover, vaccination has been associated with reduced mortality and fewer cardiovascular hospitalizations among patients with cardiovascular disease^15^. Therefore, even if NET-associated mechanisms occur rarely, the overall cardiovascular benefit–risk balance remains favorable.

These findings underscore the importance of immune–cellular interactions beyond platelets in arterial thrombosis. Experimental models of VITT and HIT (heparin-induced thrombocytopenia) highlight synergistic activation of neutrophils, monocytes, and endothelium in immune-mediated thrombosis^12^. Larger mechanistic and population-based studies are needed to clarify whether specific susceptible phenotypes exist.

## Limitations

This study cannot determine whether vaccination increases the incidence of type 1 myocardial infarction. The sample size was limited and underpowered for robust analyses by vaccine platform. As an observational study, residual confounding cannot be excluded, and causal inference is not possible.

## Conclusions

In this cohort study, SARS-CoV-2 vaccination was associated with the presence of neutrophil extracellular traps in coronary thrombi among patients with ST-segment elevation myocardial infarction. An immunothrombotic signal involving NET-mediated pathways may be implicated in post-vaccination coronary events in some patients. These findings may provide insight into thromboinflammatory processes involved in acute coronary syndromes.

## Data Availability

All data produced in the present work are contained in the manuscript

---

**Table 1.** Baseline characteristics and coronary thrombus findings among patients with STEMI according to matched SARS-CoV-2 vaccination status. Abbreviations: Anti-PF4, anti-platelet factor 4; BMI, body mass index; Cx, circumflex coronary artery; GFR (CKD-EPI), glomerular filtration rate (Chronic Kidney Disease Epidemiology Collaboration); HDL, high-density lipoprotein; hs-cTnI, high-sensitive cardiac troponin I; IQR, interquartile range; LAD, left anterior descending artery; LDL, low-density lipoprotein; N/L ratio: neutrophil-to-lymphocyte ratio; NET, neutrophil extracellular trap; NT-proBNP, N-terminal pro–B-type natriuretic peptide; PCI, percutaneous coronary intervention; PMN, polymorphonuclear; RCA, right coronary artery; SD, standard deviation; TIMI, Thrombolysis in Myocardial Infarction grade flow. ^a^TIMI grade flow: 0, no perfusion; I, penetration without perfusion; II, partial perfusion; and III, complete perfusion.

|  | SARS-CoV-2–<br>unvaccinated<br>(n=21) | SARS-CoV-2–<br>vaccinated<br>(n=23) | P value |
| --- | --- | --- | --- |

|  | **SARS-CoV-2–**<br>**unvaccinated**<br>**(n=21)** | **SARS-CoV-2–**<br>**vaccinated**<br>**(n=23)** | **P value** |
| --- | --- | --- | --- |
| **Epidemiological and clinical features** |  |  |  |
| Age, median (IQR) | 58 (49-67) | 56 (51-68) | .89 |
| Male, no. (%) | 18 (86) | 20 (87) | .91 |
| BMI, median (IQR) | 26.3 (24.2-31.8) | 25.8 (24.3-28.1) | .72 |
| Hypertension, no. (%) | 6 (29) | 8 (35) | .66 |
| Current smoker, no. (%) | 8 (38) | 11 (48) | .56 |
| Dyslipidemia, no. (%) | 11 (52) | 10 (43) | .56 |
| Diabetes, no. (%) | 3 (14) | 3 (13) | .91 |
| Ischemic cardiopathy, no. (%) | 1 (5) | 4 (17) | .19 |
| Killip status I or II, no. (%) | 20 (95) | 21 (92) | .61 |
| Time symptom-balloon (min), median (IQR) | 3.25 (2.15-5.02) | 2.83 (1.98-3.35) | .44 |
| **Angiography, no. (%)** |  |  |  |
| Infarct culprit vessel |  |  |  |
| RCA | 10 (48) | 11 (48) | .92 |
| Cx | 2 (10) | 3 (13) |  |
| LAD | 9 (43) | 9 (39) |  |
| Arterial segment, proximal | 19 (90) | 21 (92) | .53 |
| Infarction due to stent thrombosis | 1 (5) | 3 (13) | .34 |
| TIMI<sup>a</sup> status after PCI II or III | 20 (95) | 23 (100) | .29 |
| **Laboratory admission values, median**<br>**(IQR)** |  |  |  |
| Leukocyte count, cells x10<sup>3</sup>/ μ L | 12.02 (8.38-14.46) | 13.51 (9.75-15.53) | .15 |
| Lymphocyte's count, cells x10<sup>3</sup>/ μ L | 1.84 (1.40-2.60) | 1.80 (1.50-2.50) | .54 |
| N/L ratio | 3.90 (1.86-6.55) | 6.11 (4.61-8.00) | .11 |
| Hemoglobin, g/dL | 14.50 (13.90-15.50) | 15.00 (14.40-15.90) | .49 |
| Platelet count, cells x10<sup>3</sup>/ μ L | 240 (218-285) | 276 (219-329) | .28 |
| Fibrinogen, mg/dL | 316 (269-398) | 333 (275-425) | .66 |
| GFR (CKD-EPI), ml/min/1.73 m<sup>2</sup> | 0.90 (0.82-0.98) | 0.85 (0.79-1.18) | .90 |
| Peak hs-cTnI, ng/L | 274 (85-1,219) | 4,275 (114-17,863) | **.03** |
| NT-proBNP, pg/mL | 45 (10-1,028) | 239 (41.5-1,052) | .08 |
| C-Reactive Protein, mg/L | 2.9 (1.9-9.0) | 4.0 (1.6-11.8) | .60 |
| Total cholesterol, mmol/L | 176 (164-192) | 164 (127-210) | .44 |
| HDL-cholesterol, mmol/L | 40 (32-49) | 40 (33-45) | .47 |
| LDL-cholesterol, mmol/L | 116 (94-129) | 99.5 (69-120) | .19 |
| **SARS-CoV-2 positive serology, no. (%)** | 7 (33.3) | 8 (34.8) | .92 |
| IgG level, median (IQR), UA/mL | 132 (42-182) | 2081 (2015-2081) | **<0.01** |
| **Coronary thrombi analysis, no. (%)** |  |  |  |
| NETs present | 5 (24) | 14 (61) | **.01** |
| NET density, median (IQR) | 71 (58-80) | 49 (40-66) | .12 |
| PMN cells at moderate or intense levels | 9 (41) | 8 (38) | .85 |
| Fibrin, moderate or intense amount | 16 (73) | 11 (53) | .17 |
| Plaque fragments present | 10 (45) | 8 (38) | .63 |
| Anti-PF4 antibodies | 12 (57) | 7 (33) | .12 |

## REFERENCES

1. Thierry AR, Roch B. Neutrophil Extracellular Traps and By-Products Play a Key Role in COVID-19: Pathogenesis, Risk Factors, and Therapy. J Clin Med. 2020;9(9):2942. doi:10.3390/jcm9092942

2. Blasco A, Coronado MJ, Hernández-Terciado F, et al. Assessment of Neutrophil Extracellular Traps in Coronary Thrombus of a Case Series of Patients With COVID-19 and Myocardial Infarction. JAMA Cardiol. Published online December 29, 2020:1–6. doi:10.1001/jamacardio.2020.7308

3. Ferreira-da-Silva R, Lobo MF, Pereira AM, Morato M, Polónia JJ, Ribeiro-Vaz I. Network analysis of adverse event patterns following immunization with mRNA COVID-19 vaccines: real-world data from the European pharmacovigilance database EudraVigilance. Front Med. 2025;12:1501921. doi:10.3389/fmed.2025.1501921

4. Choi MG, Kim MH, Chun EM. The early impact of COVID-19 vaccines on major events in cardiac, pulmonary, and thromboembolic disease: a population-based study. Korean J Intern Med. 2025;40(5):801–812. doi:10.3904/kjim.2025.056

5. Zafar U, Zafar H, Ahmed MS, Khattak M. Link between COVID-19 vaccines and myocardial infarction. World J Clin Cases. 2022;10(28):10109–10119. doi:10.12998/wjcc.v10.i28.10109

6. Ip S, North TL, Torabi F, et al. Cohort study of cardiovascular safety of different COVID-19 vaccination doses among 46 million adults in England. Nat Commun. 2024;15(1):6085. doi:10.1038/s41467-024-49634-x

7. Xu Y, Li H, Santosa A, et al. Cardiovascular events following coronavirus disease 2019 vaccination in adults: a nationwide Swedish study. Eur Heart J. 2025;46(2):147–157. doi:10.1093/eurheartj/ehae639

8. Hippisley-Cox J, Patone M, Mei XW, et al. Risk of thrombocytopenia and thromboembolism after covid-19 vaccination and SARS-CoV-2 positive testing: self-controlled case series study. BMJ. Published online August 26, 2021:1931. doi:10.1136/bmj.n1931

9. Nitz JN, Ruprecht KK, Henjum LJ, et al. Cardiovascular Sequelae of the COVID-19 Vaccines. Cureus. Published online April 10, 2025. doi:10.7759/cureus.82041

10. Leung HHL, Perdomo J, Ahmadi Z, et al. NETosis and thrombosis in vaccine-induced immune thrombotic thrombocytopenia. Nat Commun. 2022;13(1):5206. doi:10.1038/s41467-022-32946-1

11. Santos A, MartÍn P, Blasco A, et al. NETs detection and quantification in paraffin embedded samples using confocal microscopy. Micron. 2018;114:1–7. doi:10.1016/j.micron.2018.07.002

12. Meier RT, Kapur R. Antibody-mediated multicellular pathophysiology of heparin-induced thrombocytopenia and vaccine-induced thrombotic thrombocytopenia: the dynamic roles of platelets, neutrophils, endothelial cells, and monocytes. J Thromb Haemost. 2026;24(1):4–17. doi:10.1016/j.jtha.2025.09.014

13. Kuo Y, Kang C, Lai Z, et al. Temporal changes in biomarkers of neutrophil extracellular traps and NET-promoting autoantibodies following adenovirus-vectored, mRNA, and recombinant protein COVID-19 vaccination. J Med Virol. 2024;96(3):e29556. doi:10.1002/jmv.29556

14. Fortin A, Huot S, Caron E, et al. No evidence of direct activation of human neutrophil responses by multivalent prefusion trimeric SARS-CoV-2 Spike protein ex vivo. Bonam SR, ed. PLoS One. 2025;20(10):e0332261. doi:10.1371/journal.pone.0332261

15. Akbar UA, Thyagaturu H, Taha A, et al. COVID-19 Vaccination and Cardiovascular Outcomes in Older Adults With Coronary Artery Disease and Heart Failure: Insights From a Large Propensity-Matched Cohort Study. JAHA. 2025;14(21):e044546. doi:10.1161/JAHA.125.044546

